# Population-scale genome-wide prioritization of targets for allele-specific antisense oligonucleotide therapy design

**DOI:** 10.64898/2026.10.26.26366522

**Authors:** Rebecca Sakaki, Logan Newton, Bushra Haque, Hui Lin, David Cheerie, Jacques L. Michaud, Christian R. Marshall, Philippe M. Campeau, Ashish R. Deshwar, Gregory Costain

## Abstract

**Background:** Allele-specific antisense oligonucleotides (ASOs) are increasingly used to selectively suppress transcripts containing pathogenic gain-of-function or dominant negative (GOF/DN) variants. However, most current approaches rely on patient-specific target selection of SNVs in *cis* with the disease-causing variant. This individualized approach may limit broader ASO applicability and could exacerbate inequities in access. Population-level selection of ASO-targetable SNVs is an alternative approach not previously explored at scale.

**Methods:** For a curated set of >400 disease genes with putative toxic GOF/DN mechanisms, we applied a greedy algorithm to identify ASO-targetable SNVs that maximize population coverage in the 1000 Genomes Project dataset. Population coverage was evaluated across “superpopulation” ancestry groups and assessed in multiple diverse replication datasets. For each prioritized SNV we attempted to design allele-specific 15-mer and 20-mer ASOs meeting standard chemistry and sequence criteria. The presence of prioritized SNVs was assessed in two local cohorts with GOF/DN disorders: infants with epilepsy diagnosed by rapid short-read genome sequencing, and children with long-read genome sequencing data generated for bespoke ASO development.

**Results:** We identified 7,927 SNVs (median 17.5 per gene) achieving a median cumulative heterozygous population coverage of 88.9% across 408 disease genes. More than two-thirds of genes (70.6%) reached ≥80% coverage using a median of only four prioritized SNVs. Coverage was consistently high for all “superpopulation” ancestry groups, with median coverage proportions of ≥85%. Prioritized SNVs generalized to independent ancestrally diverse replication datasets, achieving median coverage of 86.4% in the Human Genome Diversity Project and 90.3% in the *All of Us* Research Program. Many prioritized targets were compatible with established ASO design principles, enabling generation of candidate allele-specific 15-mer and 20-mer ASOs at scale for future experimental validation. The presence of prioritized targets in the rapid short-read sequencing cohort, and the overlap between prioritized targets and manually curated long-read genome sequencing targets, supports the clinical relevance of population-informed target selection. All SNV targets and candidate allele-specific ASO sequences are available at hawk-eye.research.sickkids.ca/knockdown.

**Conclusions:** Prioritization of common SNV targets may facilitate the development of ASO therapies with broader population coverage, helping move the field beyond single-patient applications and toward more equitable opportunities for access. Target prioritization for allele-specific antisense oligonucleotides

## BACKGROUND

Antisense oligonucleotides (ASOs) are a versatile class of RNA therapeutics for inborn genetic diseases. An ASO may be DNA variant-agnostic or variant-specific and can even represent a truly individualized genetic intervention for a novel but uniquely targetable DNA variant (1–6). Proofs-of-concept exist for allele-specific downregulation of transcripts arising from toxic gain-of-function (GOF) or dominant negative (DN) variants, such as variants causing *KIF1A* related neurological disorder (MONDO:0700055) (7) or *SCN2A* developmental and epileptic encephalopathy (MONDO:0013388) (8,9). The predominant strategy in the field has been to develop an allele-selective knockdown ASO based on a single individual’s genetic profile, often by targeting a heterozygous single-nucleotide variant (SNV) in *cis* with the disease-causing variant, for use in a single-patient trial (1,7,10). The resulting ASO is more likely to be uniquely applicable to the patient of interest or a small subgroup of individuals, rather than broadly applicable across the disease population. This paradigm may inadvertently constrain scalability and (if the target SNV has variable allele frequency globally) compound genetic ancestry-mediated disparities in research opportunities and treatment. In contrast, deliberate population-level prioritization of target SNVs for allele-specific ASO-mediated knockdown may help mitigate initial systematic inequities in access to experimental genetic therapies. This concept was recently explored retrospectively for *KIF1A* related neurological disorder (11). We hypothesized that such an approach could be scaled to all known autosomal dominant disease genes with GOF/DN mechanisms.

In this study, we applied a greedy algorithm to an ancestry-diverse catalogue of human genetic variation to identify common, ASO-targetable SNVs that maximize population coverage globally, for each of the >400 disease genes associated with GOF/DN disorders. We further demonstrated that *in silico*-optimized ASO sequences can be designed at scale for these targets, and showed feasibility of this *a priori* target selection approach using long-read genome sequencing data from a bespoke ASO development program. All SNV targets and candidate allele-specific ASO sequences are made available in HAWK-EYE, an open-access searchable web application (12).

## METHODS

### Autosomal dominant gain-of-function and dominant negative gene list

Genes associated with AD GOF/DN disorders were curated from multiple sources: (i) 235 genes were obtained from Gene2Phenotype (13) based on filtering by allelic requirement (monoallelic_autosomal OR monoallelic_X_heterozygous), variant consequence (altered gene product OR increased gene product level), confidence (definitive OR strong OR moderate) and molecular mechanism (gain-of-function OR dominant negative OR activating); (ii) 212 genes were obtained from the LoGoFunc’s Human Gene Mutation Database (14) and ClinVar (15) databases by filtering for a GOF mechanism; (iii) an additional 36 genes were added via literature review. The union was 409 unique genes (Supplemental Table 1). Coordinates for MANE Select transcripts were retrieved from GENCODE (Release 49, GRCh38) (16).

### Common SNV extraction and filtering in the discovery dataset

1000 Genomes Project (1kGP) phase 3 short-read genome sequencing variant calls were obtained for 3,202 individuals in Variant Call Format (VCF) from five “super populations”: African (AFR), Admixed American (AMR), East Asian (EAS), European (EUR), and South Asian (SAS). To minimize relatedness within the dataset, offspring from trios were excluded, resulting in a dataset of 2,594 directly unrelated individuals. Common SNVs (minor allele frequency >1% across the aggregate 1kGP dataset) were extracted for each of the 409 GOF/DN genes in our list across participants. To filter out SNVs in suboptimal regions for ASO-based approaches (Alu elements, tandem repeats), we downloaded RepeatMasker and Tandem Repeats Finder annotations from the UCSC genome annotation database for the Dec. 2013 (GRCh38/hg38) assembly of the human genome (17). Common SNVs within these regions were excluded from further analyses.

### Greedy algorithm for SNV prioritization

A greedy algorithm was applied to the extracted 1kGP common SNVs to prioritize SNVs that maximize cumulative heterozygous population coverage. The algorithm identifies heterozygous participants for each common SNV and iteratively selects SNVs per gene that capture the largest number of “uncovered” heterozygous participants (those not heterozygous for any of the previously selected SNVs). Selection continues until either (i) ≥95% coverage or (ii) saturation, with no additional SNVs contributing to increased coverage.

### Linkage disequilibrium analysis

Linkage disequilibrium (LD) between pairs of prioritized SNVs was calculated using PLINK 2.0 (version a5.12) (18,19). VCF files were first filtered for prioritized SNVs separated by superpopulation group and were converted to pgen format using the --make-pgen function of PLINK 2.0. Pairwise LD statistics (r^2^ and D’) were computed independently for each superpopulation group using the --r2-phased function (ref-based cols=+dprime). Variant pairs were restricted to those with r^2^ ≥ 0.8 and positive D’ to capture SNV pairs for which the alternate alleles were in strong linkage disequilibrium with each other.

### Superpopulation-specific coverage of prioritized SNVs

Per-gene coverage of the prioritized SNVs was calculated separately for each 1kGP superpopulation (AFR, AMR, EAS, EUR, SAS). For each gene, coverage was defined as the proportion of individuals heterozygous for at least one SNV in our prioritized set. The coverage median and interquartile range were calculated across all 408 genes within each superpopulation.

### Defining two independent replication datasets

Two additional short-read genome sequencing variant call datasets were obtained from the Human Genome Diversity Project (HGDP) and the *All of Us* Research Program. The HGDP comprised 780 unrelated samples from the publicly available gnomAD v3 HGDP and 1kGP callset (20,21). Samples were restricted to individuals included exclusively in HGDP and not represented in 1kGP. The *All of Us* Research Program comprised 492,799 individuals post-filtering for unrelated individuals with predicted ancestry groups (22). The coverage median and interquartile range were calculated across all 408 genes across and within each superpopulation.

### Additional aggregate allele frequency datasets

We searched for additional open-access databases of subpopulation genetic variation found by genome sequencing (i.e., including intronic regions). Aggregate allele frequencies were obtained from the Turkish Variome and ABraOM, representing Turkish people from Turkey, and Brazilian people from Brazil, respectively (23,24). The Turkish Variome comprises variant calls from 3,362 exome or genome sequencing samples and ABraOM comprises variant calls from 1,171 genome sequencing samples. We filtered variants for overlap with our prioritized SNV set and calculated the proportion of individuals that were heterozygous for each overlapping SNV. For the Turkish Variome, exonic variants contained ∼4.5x more genotyped alleles due to the presence of some exome-only sequencing data.

### Disease phenotype classification

Gene-disease subgroup associations were obtained from Genomics England PanelApp by downloading curated gene panels annotated with relevant Level 2 disease subgroups (download date: April 2026) (25). For each disease subgroup, the corresponding gene set was compiled and intersected with the High and Moderate Feasibility gene groups.

### Allele-specific knockdown ASO design

We developed a Python-based tool to generate and prioritize allele-specific 15-mer and 20-mer knockdown ASO sequences. This tool accepts as input a target DNA sequence containing the prioritized SNV with a minimum of 250 bp flanking sequence on each side. All possible 15-mer and 20-mer sequences are generated by tiling across the input sequence, and only those overlapping the target SNV are retained. The ASO sequences are then filtered in accordance with established ASO design principles with the incorporation of additional computational tools (26). Off-target filtering uses BLAST against RefSeqGene sequences to exclude any candidate sequence with 15/15 bp (15-mers) or ≥17/20 bp (20-mers) consecutive identity to a non-target sequence. A full description of incorporated tools and criteria is provided elsewhere (27).

### ClinVar and common variation nearby prioritized SNVs

Common SNVs within +/-14 bp and within +/-19 bp of each prioritized SNV were identified from the 1kGP and gnomAD v4.1 (joint exome-genome; PASS variants; allele frequency >1%) (28), restricted to gene boundaries defined by the MANE Select transcript coordinates used for common SNV extraction and filtering. ClinVar variant data were retrieved from the publicly available clinvar.vcf.gz file (file date: June 6, 2026) (15) and filtered for missense germline pathogenic/likely pathogenic SNVs with at least a two-star review status, within genes in our list. These variants serve as a proxy for recurring GOF or DN variants causing AD diseases in our genes of interest. Filtered ClinVar variants within +/-14 bp or +/-20 bp of each prioritized SNV were identified.

### SNV analysis from short- and long-read genome sequencing data

Genome sequencing for participants in the Gene-STEPS study was performed using short-read Illumina platforms and published methods (3,29,30). Long-read genome sequencing for bespoke ASO development was performed via PacBio Revio SMRT cell sequencing experiments using high-quality blood-derived genomic DNA. Sequencing, alignment, and variant calling was performed using standard methods at The Centre for Applied Genomics (TCAG; Toronto, Canada). Ancestry information was obtained through parent self-report during family history collection. For analytical purposes, self-reported ancestry categories were mapped to the broad superpopulation groupings used in the genomic analyses wherever possible. To protect participant anonymity, individual-level ancestry categories and DNA variants are not shown.

## RESULTS

### Prioritized common heterozygous SNVs achieve high population coverage across GOF/DN genes

To identify a minimal set of common heterozygous SNVs that could enable broad applicability of allele-specific ASO knockdown strategies, we leveraged the variant call dataset from the 1kGP and applied the prioritization framework outlined in Fig. 1. First, we extracted common (minor allele frequency >1%) exonic and intronic SNVs from the aggregate 1kGP dataset across 409 autosomal dominant disease genes with putative or established GOF/DN mechanisms (Supplemental Table 1). After filtering out SNVs in suboptimal regions for ASO-based approaches (see Methods), 126,829 SNVs remained [per gene: mean 310.1, median 113 (interquartile range (IQR) 36-319)]. One gene (*SIK1*) had no common SNVs and was excluded from further analyses. We then applied a greedy algorithm to prioritize extracted SNVs that maximize cumulative heterozygous 1kGP population coverage (hereafter coverage refers to heterozygous coverage). The algorithm output a total of 7,927 unique SNVs across 408 genes [per gene: mean 19.4, median 17.5 (IQR 8.8-27.0)] (Fig. 2a, Supplemental Table 2).

**Fig. 1:**
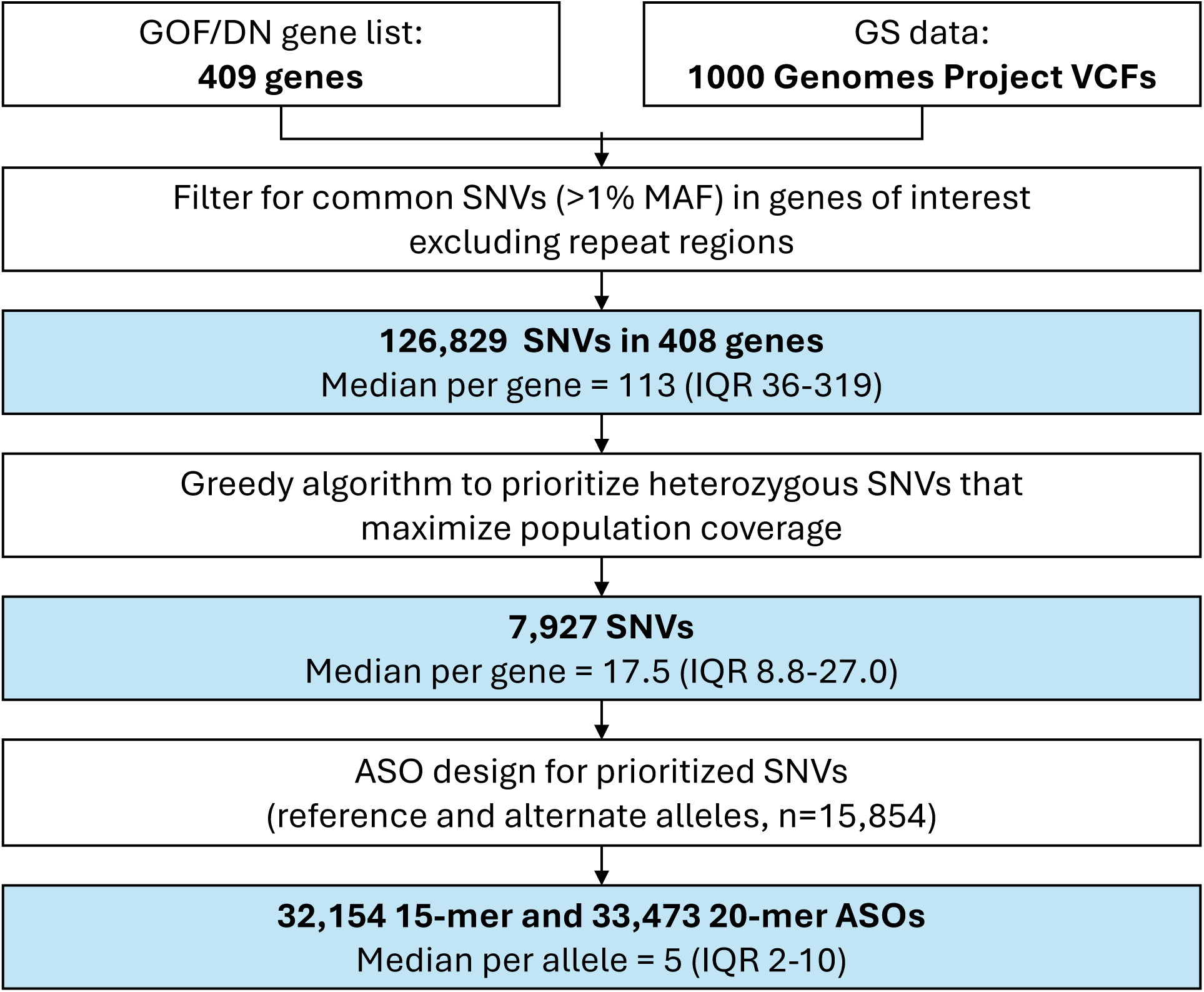
Greedy prioritization of ASO-targetable SNVs for population coverage. Study overview. Autosomal dominant disease genes with putative or established gain-of-function or dominant negative (GOF/DN) mechanisms were compiled from Gene2Phenotype, LoGoFunc’s Human Gene Mutation Database, ClinVar, and a literature review. Common single-nucleotide variants (SNVs) from the 1000 Genomes Project genome sequencing (GS) Variant Call Format (VCF) files were used as input to a greedy algorithm to prioritize SNVs for antisense oligonucleotide (ASO) design. MAF, minor allele frequency.

**Fig. 2:**
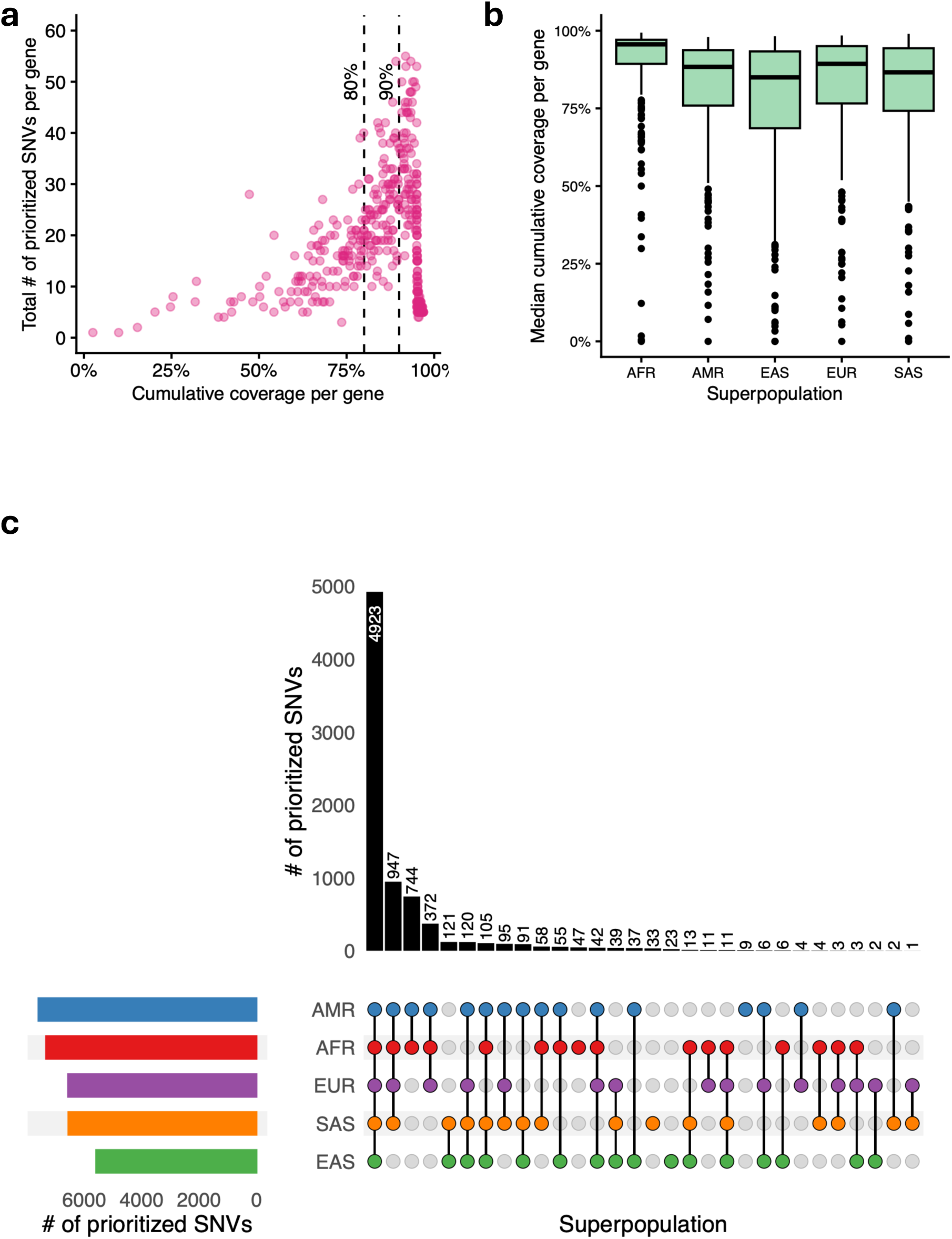
Population coverage of prioritized SNVs across 1kGP superpopulations. **a** Scatterplot showing cumulative heterozygous population coverage achieved per gene with our prioritized SNVs versus the total number of prioritized SNVs per gene. Each point represents a single gene. **b** Boxplot showing the distribution of cumulative heterozygous population coverage across the 1000 Genomes Project superpopulations. Boxes represent the interquartile range (IQR), the central line indicates the median, whiskers extend to observations within 1.5 × the IQR and points represent outliers. AFR, African; AMR, Admixed American; EAS, East Asian; EUR, European; SAS, South Asian. **c** UpSet plot showing overlap of prioritized SNVs observed in a heterozygous state across the five 1kGP superpopulations. Bars represent the number of prioritized SNVs observed in a heterozygous state in at least one individual within the indicated superpopulation groups. Set size bars represent the total number of prioritized SNVs observed in a heterozygous state in at least one individual within each superpopulation.

Overall, 70.6% (288/408), 46.8% (191/408), and 31.1% (127/408) of genes achieved ≥80%, ≥90%, and ≥95% cumulative population coverage, respectively. The median minimum number of SNVs required per gene to reach these coverage thresholds was four, six, and seven, respectively. Put differently, for each of 288 genes associated with GOF/DN diseases, ASOs targeting a median of four different predetermined SNV targets would be sufficient to treat ≥80% of individuals represented by the discovery population dataset. Across all genes, the median cumulative coverage was 88.9% (IQR 78.0-95.1%). Although additional prioritized SNVs increased cumulative coverage, only a small number of SNVs were required to cover most individuals.

Prioritized SNVs may be redundant for ASO design if they are in strong LD, as individuals with multiple SNVs within the same haplotype block could potentially be treated by a single ASO targeting any one of those SNVs. To assess redundancy amongst prioritized SNVs, we calculated the proportion of prioritized SNVs that are in high LD (r² > 0.8) with at least one other prioritized SNV. Given known differences in haplotype structure across genetic ancestry groups, LD was assessed separately within each 1kGP superpopulation. Only 6.1% (483/7,927) of prioritized SNVs exhibited high LD with another across all superpopulations (Supplemental Table 3), suggesting largely independent contributions of prioritized SNVs to population coverage.

### Prioritized SNVs achieve high coverage across superpopulations

To explore potential ancestry bias, we assessed superpopulation-specific coverage. Across 408 genes, the median coverage proportions were ≥85% for all superpopulations [AFR: 95.6% (IQR 89.6-97.1%), AMR: 88.4% (75.9-93.8%), EAS: 85.0% (68.6-93.4%), EUR: 89.4% (76.6-95.1%), SAS: 86.6% (74.2-94.4%)] (Fig. 2b, Supplemental Table 4). Prioritized SNVs were largely shared between superpopulations. Most SNVs were shared across multiple superpopulations, with 62.1% (4,923/7,927) observed in a heterozygous state across all five superpopulations and 86.8% (6,883/7,927) observed in at least three superpopulations (Fig. 2c). This substantial overlap indicates our set of SNVs can provide coverage across diverse genetic ancestries.

### Prioritized SNVs generalize to independent, ancestrally diverse variant-call datasets

To assess the generalizability of our prioritized SNV set, we evaluated cumulative per-gene coverage in two independent short-read genome sequencing variant call datasets from the HGDP and the *All of Us* Research Program. These datasets comprise individuals of AFR, AMR, EAS, Middle Eastern (MID), EUR, and SAS ancestry (21,22). Across all 408 genes, prioritized SNVs achieved a median coverage of 86.4% (IQR 73.8-92.3%) in the HGDP cohort and 90.3% (IQR 78.4-95.0%) in the *All of Us* cohort (Supplemental Fig. 1). Median coverage was 2.5 percentage points lower in HGDP and 1.4 percentage points higher in *All of Us* compared to 1kGP, despite the broader range of geographically diverse populations and thousands more individuals represented in these datasets.

Many population groups are not represented in either the 1kGP, HGDP, or *All of Us* datasets. To show that our approach is relevant to additional populations, we leveraged publicly accessible aggregated allele frequencies from genome-wide sequencing of Turkish and Brazilian cohorts. Both cohorts are unrelated, highly admixed populations and represent genetic ancestry across multiple superpopulation groups. Among prioritized SNVs, 86.0% (6,818/7,927) and 98.9% (7,843/7,927) were observed in the Turkish and Brazilian cohorts, respectively. Most genes [92.9% (379/408) in Turkish cohort, 95.1% (388/408) in Brazilian cohort] had at least one prioritized SNV that was heterozygous in >25% of the population, representing a substantial proportion of possible heterozygotes considering the theoretical maximum frequency of 50%.

### Most GOF/DN genes are amenable to broadly applicable ASO design

We defined four mutually exclusive gene categories to describe the prospect of developing a small number of ASOs to treat most patients with the associated disease:

i. High Feasibility: ≥90% population coverage with ≤10 SNVs;
ii. Moderate Feasibility: ≥80% to <90% coverage with ≤10 SNVs;
iii. Limited Feasibility: ≥80% coverage only with >10 SNVs;
iv. Low Feasibility: <80% coverage regardless of SNV number.

There were 140 genes (34.3%) in the High Feasibility group and 124 genes (30.4%) in the Moderate Feasibility group (Fig. 3a, Supplemental Table 5). Substantial coverage is achieved for High and Moderate Feasibility genes with a small number of SNVs (Fig. 3b). Among these 264 genes, 200 (75.8%) were associated with at least one of developmental, neurological, or neurological and neurodevelopmental disorders in Genomics England PanelApp, which are major areas of current ASO therapeutic interest (Supplemental Table 1).

**Fig. 3:**
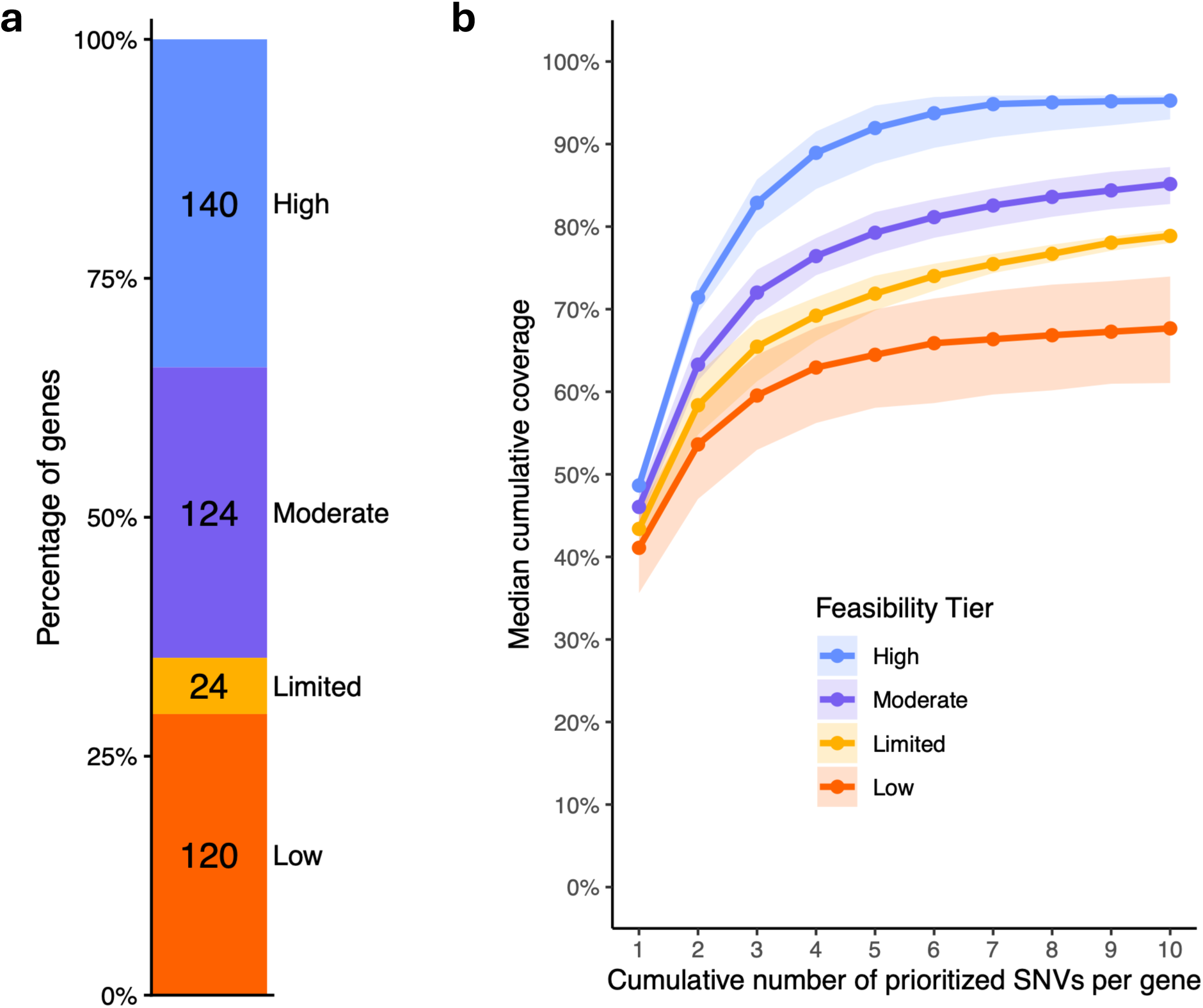
Distribution of gene feasibility tiers and coverage achieved by prioritized SNVs. **a** Proportion and number of genes classified into mutually exclusive categories to describe the feasibility of developing a small number of ASOs to treat most patients with the associated disease. **b** Median cumulative heterozygous population coverage achieved by prioritized SNVs per gene, stratified by feasibility tier. Shaded regions represent the interquartile range.

### Greedy algorithm prioritization identifies broader-coverage SNV targets compared with ranking by heterozygote frequency alone

To evaluate the benefit of the greedy algorithm approach, we compared our prioritized SNVs with a simpler strategy in which ten SNVs with the highest heterozygote frequencies were selected from the 1kGP variant dataset post-filtering for repeat regions (Supplemental Table 6). We calculated the cumulative population coverage per gene using these ten SNVs and assigned genes to our feasibility categories. Across all genes, the median cumulative coverage was 71.6% (IQR 61.4-82.5%), representing a 17.3-percentage-point decrease in median coverage compared with the greedy approach. In addition, the discrepancies between superpopulation coverage increased, with a 15.5% difference between the lowest covered (EAS, median coverage: 65.8%) and highest covered (AFR, median coverage: 81.3%) superpopulations, compared to a 10.6% difference with our greedy approach. Using the frequency-based strategy, 55 genes (13.5%) were classified as High Feasibility, while 69 (16.9%) genes were classified as Moderate Feasibility. Greedy prioritization increased the number of genes classified as High or Moderate Feasibility from 124 (30.4%) to 264 (64.7%), representing a 2.1-fold improvement. Notably, all 124 genes identified as High or Moderate Feasibility using the frequency-based strategy were also recovered by the greedy prioritization. Of these, 44 (35.5%) were upgraded from Moderate to High Feasibility using greedy prioritization, whereas no genes were downgraded (Supplemental Table 5).

### Most prioritized SNV targets have regional properties consistent with established ASO design principles

We explored if SNVs were generally suitable for ASO targeting, by attempting to design 15-mer and 20-mer sequences at scale that would meet established sequence-, chemical-, and thermodynamic properties. Typically, knockdown ASOs are 20 bp in length; however, shorter ASOs may provide greater sensitivity to single-nucleotide mismatches and improved allele-selectivity (31). As the phase between pathogenic (typically *de novo*) variants and prioritized SNVs is often initially unknown due to the widespread use of short-read sequencing, we proactively designed ASO sequences against both reference and alternate alleles, at each prioritized SNV position (7,927×2, n=15,854). Following filtering based on desired sequence properties, 32,154 15-mer and 33,473 20-mer candidate ASOs were retained. A majority (58.7%; 9,285/15,854) of prioritized SNV alleles yielded at least one candidate *in silico*-optimized, allele-specific ASO. A median of 5 (IQR 2-10) candidate ASOs were designed per SNV allele, and 46.3% (3,671/7,927) of SNV positions had at least one candidate ASO for both the reference and alternate alleles. All ASO sequences passing filtering are available on HAWK-EYE as candidate designs for further optimization and experimental validation (12).

Nearby sequence variants can compromise ASO binding if they introduce mismatches relative to the reference genome. We identified common SNVs from 1kGP and gnomAD v4.1 datasets that were located +/-14 bp upstream or downstream of the prioritized SNVs and could potentially interfere with 15-mer ASO binding. Only 13.3% and 12.2% of prioritized SNVs had at least one common SNV in 1kGP and gnomAD v4.1 in this window, respectively. GOF/DN disorders can be caused in unrelated individuals by recurring pathogenic variants, which could limit ASO use if the pathogenic variant is commonly within the ASO binding region of a prioritized SNV. Recurrent germline pathogenic or likely pathogenic missense variants in ClinVar were used as a proxy for these pathogenic variants, resulting in only 0.4% of prioritized SNVs having a ClinVar variant +/-14 bp upstream or downstream. Results were similar when considering a +/-19 bp window for 20-mer ASOs: 16.4% and 15.1% of prioritized SNVs had at least one common SNV in 1kGP and gnomAD v4.1, respectively, and 0.5% had a ClinVar variant.

### Clinically relevant SNV targets are identified by our approach

In a local research cohort of neonates and infants who underwent rapid short-read genome sequencing for epilepsy, we identified 15 participants with *de novo* variants causing 11 different GOF/DN disorders (disease-associated genes: *ATXN2*, *BRAF*, *DEAF1*, *FGF12*, *GRIN1*, *KCNQ2*, *KCNT1*, *RHOBTB2*, *SCN1A*, *SCN2A*, and *SCN8A*). The self-reported ancestries of 27/30 parents mapped to 1kGP superpopulations: EUR (n=11), SAS (n=8), AFR (n=6), EAS (n=2). Overall, 14/15 participants were heterozygous for at least one SNV identified by our greedy algorithm in the relevant gene. The participant without prioritized SNVs had a reported ancestry known to be underrepresented in all population datasets.

For five additional individuals with GOF/DN disorders enrolled in a local bespoke ASO development program, phased PacBio long-read genome sequencing data were previously used to identify heterozygous SNVs in *cis* with the *de novo* disease-causing variant. At least four of these SNVs per patient were independently prioritized by our greedy algorithm approach, demonstrating overlap between empirically identified, manually curated patient-specific targets and population-informed target selection (Table 1). For every patient, at least one overlapping SNV target within the affected gene ranked within the top five prioritized SNVs by added population coverage, suggesting that clinically actionable targets identified in this work have potential for use at the disease-community level. Further, all patients had at least three ASO sequences targeting the overlapping SNVs that pass our ASO filtering criteria.

**Table 1:**
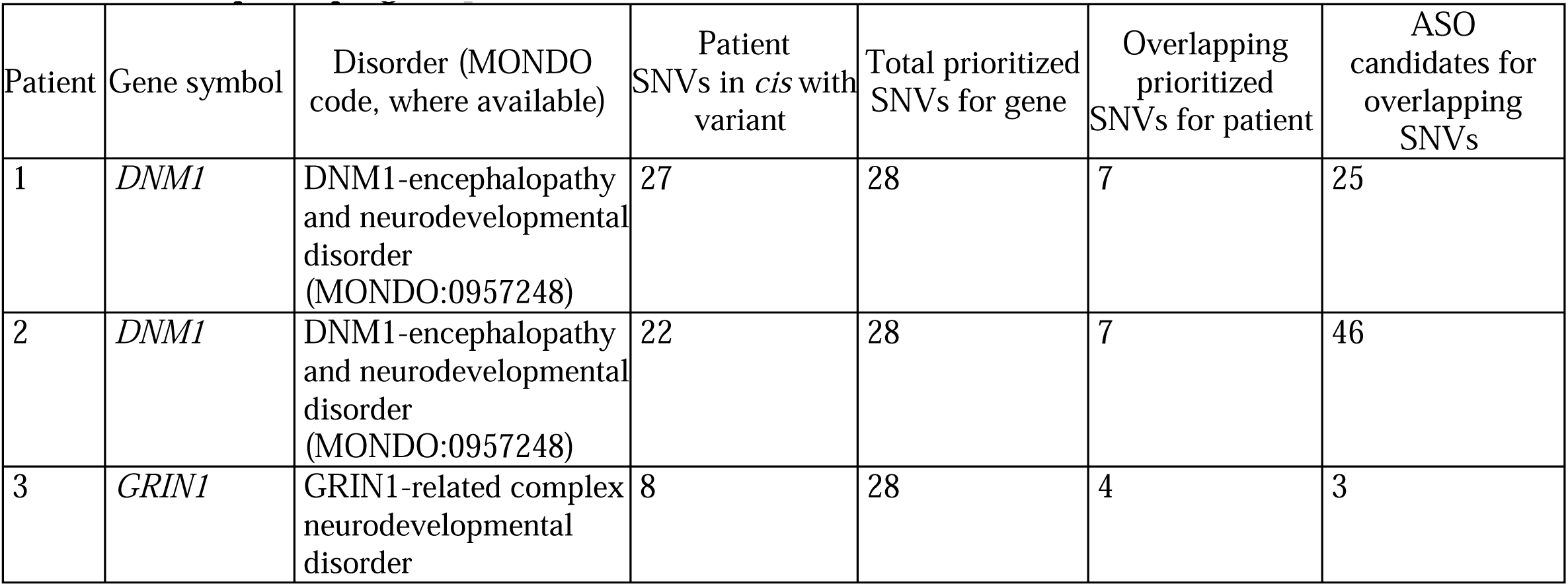
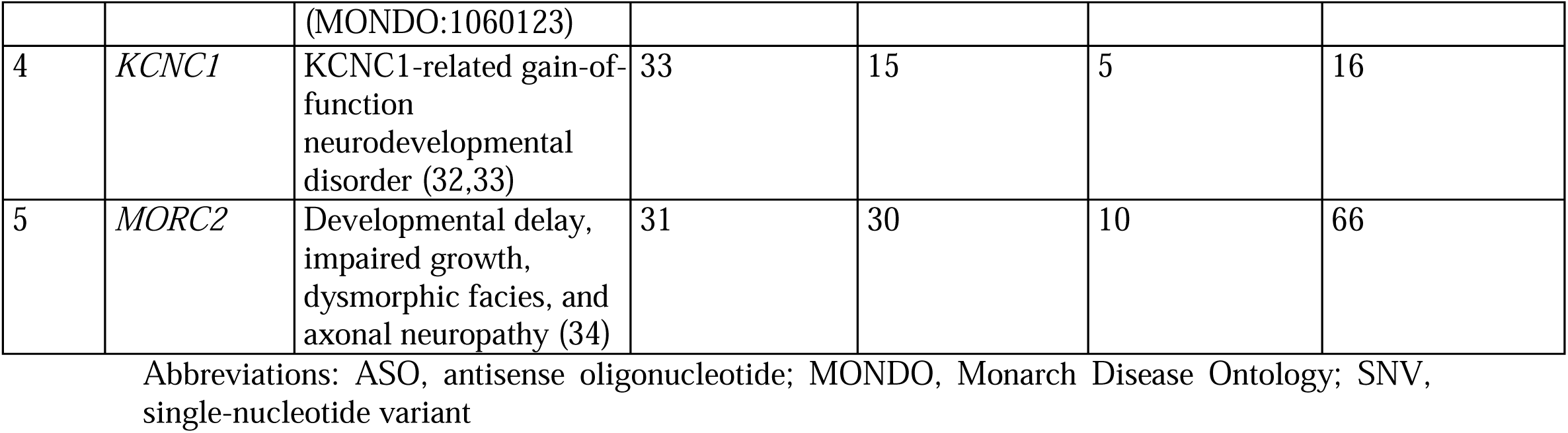
Overlap of prioritized SNVs with patient-specific targets from a bespoke ASO development program.

## DISCUSSION

Allele-selective knockdown ASOs are an emerging therapeutic strategy for toxic GOF or DN variants underlying rare and ultra-rare diseases. However, selecting the target sequence for these ASOs based on a single individual’s genetic profile may unnecessarily limit the applicability to others in the disease community, and risks exacerbating therapeutic and research inequities connected (directly or indirectly) to genetic ancestry (35). We applied a greedy algorithm to an ancestrally diverse human variant dataset and identified common heterozygous SNVs across 408 genes with putative GOF or DN mechanisms that represent robust targets for allele-selective knockdown ASOs across populations. We identified a substantial subset of genes where most individuals with the associated GOF/DN disease could be treated with a small number of allele-selective ASOs (Moderate and High feasibility). That is, for many genes, broad population coverage could be achieved through targeting a modest number of common SNVs. We replicated the primary findings using additional large datasets. We also provided preliminary evidence that our greedy algorithm approach outperforms simpler alternatives, that the prioritized SNV targets are therapeutically tractable (not just statistically common), and that findings are relevant to real-world patient cohorts.

Realizing the full potential of interventional genomics will require innovation at all stages from development to trials to clinical implementation (36). In the context of customizable nucleic acid therapeutics such as ASOs, considerations of broader applicability during early preclinical development may increase the potential health benefit for a substantially larger number of individuals within a disease community. This study illustrates secondary benefits beyond improved genome diagnostics of investing in the creation of diverse genomic variant datasets. Aligning with calls to maximize learning and knowledge sharing across rare disease therapeutic development programs, population-scale target selection may offer additional benefits beyond increasing patient coverage. By identifying recurring, ancestry-diverse candidate SNVs, this approach may enable opportunities for sharing of preclinical findings, biological insights, ASO design experience, and safety data that may have relevance beyond a single patient and can potentially inform subsequent genetic therapy development efforts. In a future state, we anticipate the HAWK-EYE web application to catalogue such information alongside prioritized candidate SNVs. Requests to contribute to HAWK-EYE entries based on published or unpublished data germane to prioritized candidate SNVs can be submitted via the e-mail contact on the HAWK-EYE website (12).

This work is a step towards more equitable design of genetic therapies. However, health disparities in access to genetic medicines exist across the entire therapeutic pipeline, including diagnosis, therapy development, regulatory pathways, manufacturing capacities, and therapeutic delivery (35,37,38). Addressing these inequities is a necessary step that requires international collaboration and intentional consideration of scientific, ethical, legal and societal factors that influence access to emerging genetic therapies (35,39). Our approach has other limitations. The 1kGP “superpopulation” groups do not fully reflect global genetic diversity and may not capture ancestry-or community-specific variation relevant to clinical implementation (40,41). Observed ancestry-associated differences in coverage at the “superpopulation” level further underscore the value of culturally informed, community-supported efforts to generate sequence data for under-represented subgroups (41). Recent initiatives have demonstrated the utility of such data in supporting precision medicine implementation (42). There is residual uncertainty for many genes regarding GOF/DN pathomechanisms, and our framework is readily adaptable as disease-gene knowledge evolves. We acknowledge that ASO therapies are not well suited for all diseases and affected organs. However, intrathecal ASO delivery has shown promise for treating neurological and neurodevelopmental disorders (5,43), one of the most represented disease categories. ASO candidates were prioritized *in silico* as a secondary finding supporting the potential suitability of prioritized SNVs as targets and are not intended to serve as strong pre-clinical design leads without further optimization. We also emphasize that ASOs require experimental validation for potency, allele specificity, toxicity, and delivery to disease-relevant tissues. Although no single allele-specific ASO is expected to be applicable to an entire disease community, this approach identifies a limited set of ASOs to collectively maximize coverage.

## CONCLUSIONS

In summary, we show our prioritized SNVs are broadly applicable across multi-ethnic genomic datasets and specific subpopulation groups. We identified at least 264 genes where thoughtful proactive SNV target selection for allele-specific ASO design could greatly benefit the global disease community. Prioritizing common SNVs with high population coverage may aid in extending the benefit of ASO therapies beyond single-patient applications and support ongoing efforts to develop treatment options for rare and ultra-rare genetic diseases at scale.

## LIST OF ABBREVIATIONS

AD: Autosomal dominant
AFR: African
AMR: Admixed American
ASO: Antisense oligonucleotide
DN: Dominant negative
EAS: East Asian
EUR: European
GOF: Gain-of-function
HGDP: Human Genome Diversity Project
IQR: Interquartile range
LD: Linkage disequilibrium
SAS: South Asian
SNV: Single-nucleotide variant
VCF: Variant Call Format
1kGP: 1000 Genomes Project

## DECLARATIONS

### Ethics approval and consent to participate

Participants with short-read or long-read genome sequencing data were recruited and provided written informed consent under research studies approved by the Research Ethics Board at the Hospital for Sick Children.

### Consent for publication

Not applicable.

### Availability of data and materials

The 1000 Genomes Project and Human Genome Diversity Project datasets analyzed during the current study are available to download from gnomAD (44). This study used data from the *All of Us* Research Program’s Controlled Tier Dataset [version 9], available to authorized users on the Researcher Workbench. Phased long-read genome sequencing data for individuals from our ASO development program will be made available through the HiFi Solves Consortium. All SNV targets and candidate ASO sequences are available through the HAWK-EYE web application at hawk-eye.research.sickkids.ca/knockdown. All code generated for this work will be available on our GitHub (45).

## Authors’ contributions

Conceptualization: GC

Data curation: RS, LN, HL, BH, DC

Formal analysis: RS, LN

Funding acquisition: JM, CRM, PC, ARD, GC

Supervision: JM, CRM, PC, ARD, GC

Visualization: RS

Writing-original draft: RS, LN, GC

Writing-review & editing: HL, BH, DC, JM, CRM, PC, ARD

## Data Availability

The 1000 Genomes Project and Human Genome Diversity Project datasets analyzed during the current study are available to download from gnomAD. This study used data from the *All of Us Research Programs* Controlled Tier Dataset [version 9], available to authorized users on the Researcher Workbench. Phased long-read genome sequencing data for individuals from our ASO development program will be made available through the HiFi Solves Consortium. All SNV targets and candidate ASO sequences are available through the HAWK-EYE web application at hawk-eye.research.sickkids.ca/knockdown. All code generated for this work will be available on our GitHub.

https://hawk-eye.research.sickkids.ca/

https://github.com/CostainLab/knockdown_aso

## Acknowledgements

We gratefully acknowledge *All of Us* participants for their contributions, without whom this research would not have been possible. We also thank the National Institutes of Health’s *All of Us* Research Program for making available the participant data examined in this study. We thank the larger Gene-STEPS and DECODE(U)R study teams, and all the patients and families who contribute to these large-scale multi-site research projects. Data processing and data analysis were performed at the High-Performance Computing Facility, Centre for Computational Medicine, The Hospital for Sick Children, Toronto, Canada.

## Supplemental Files

Supplemental Material 1: Contains Supplemental Tables 1-6 (.xlsx file).

Supplemental Material 2: Contains Supplemental Fig. 1 (.pdf file).

## REFERENCES

1. Cheerie D, Meserve MM, Beijer D, Kaiwar C, Newton L, Taylor Tavares AL, et al. Consensus guidelines for assessing eligibility of pathogenic DNA variants for antisense oligonucleotide treatments. The American Journal of Human Genetics. 2025 May;112(5):975–83. doi:10.1016/j.ajhg.2025.02.017

2. Cheerie D, Lauffer MC, Newton L, Amburgey K, Beijer D, Haque B, et al. Screening rare genetic diagnoses for amenability to bespoke antisense oligonucleotide therapy development: A retrospective cohort study. Genetics in Medicine. 2026 Jan 1;28(1):101597. doi:10.1016/j.gim.2025.101597

3. Sherrill E, Cheerie D, Beck CJ, Whittle EF, Shafi Y, Chandler NJ, et al. Precision Antisense Oligonucleotide Therapy Amenability for Infantile Genetic Epilepsies. JAMA Neurol. 2026 Jun 1;83(6):598–601. doi:10.1001/jamaneurol.2026.1021

4. Nakayama T, El Achkar CM, Burbano LE, Quraishi IH, Wu J, Li M, et al. Antisense oligonucleotide-mediated knockdown therapy in two infants with severe KCNT1 epileptic encephalopathy. Nat Med. 2026 Apr;32(4):1411–20. doi:10.1038/s41591-026-04314-9

5. Miller TM, Cudkowicz ME, Genge A, Shaw PJ, Sobue G, Bucelli RC, et al. Trial of Antisense Oligonucleotide Tofersen for SOD1 ALS. New England Journal of Medicine. 2022 Sep 21;387(12):1099–110. doi:10.1056/NEJMoa2204705

6. Ahonen S, Nitschke S, Grossman TR, Kordasiewicz H, Wang P, Zhao X, et al. Gys1 antisense therapy rescues neuropathological bases of murine Lafora disease. Brain. 2021 May 16;144(10):2985–93. doi:10.1093/brain/awab194 PubMed PMID: 33993268; PubMed Central PMCID: PMC8634080.

7. Ziegler A, Carroll J, Bain JM, Sands TT, Fee RJ, Uher D, et al. Antisense oligonucleotide therapy in an individual with KIF1A-associated neurological disorder. Nat Med. 2024 Oct;30(10):2782–6. doi:10.1038/s41591-024-03197-y

8. Kim-McManus O, Mignon L, Douville J, Pu H, Parisien C, Glass S, et al. Individualized antisense oligonucleotides for SCN2A-related developmental epileptic encephalopathy. Nat Med. 2026 Aug;32(8):2838–45. doi:10.1038/s41591-026-04527-y

9. Wagner M, Berecki G, Fazeli W, Nussbaum C, Flemmer AW, Frizzo S, et al. Antisense oligonucleotide treatment in a preterm infant with early-onset SCN2A developmental and epileptic encephalopathy. Nat Med. 2025 Jul;31(7):2174–8. doi:10.1038/s41591-025-03656-0

10. Crooke ST, Glass S, Gleeson JG, Mignon L, Skourti-Stathaki K, Douville J, et al. Addressing the needs of nano-rare patients: the n-Lorem experience. Nucleic Acids Res. 2026 Jun 10;54(10):gkag504. doi:10.1093/nar/gkag504

11. Zuccaro MV, Young RE, Hu J, Lanzano P, Semenova E, Lin X, et al. Antisense oligonucleotides to KIF1A polymorphisms expand targets and rescue patient-derived neurons in vitro. Nat Commun. 2026 Jan 9;17(1):1109. doi:10.1038/s41467-025-67752-y

12. HAWK-EYE [Internet]. The Hospital for Sick Children. HAWK-EYE. Available from: https://hawk-eye.research.sickkids.ca/

13. Hunt SE, Lemos D, Pericherla SR, Austine-Orimoloye O, Cibrian Uhalte E, Yates TM, et al. Gene2Phenotype: A Database of Structured Human Monogenic Diseases and Pathomechanisms. Journal of Molecular Biology. 2026 Mar 26;169768. doi:10.1016/j.jmb.2026.169768

14. Bayrak CS, Stein D, Jain A, Chaudhary K, Nadkarni GN, Vleck TTV, et al. Identification of discriminative gene-level and protein-level features associated with pathogenic gain-of-function and loss-of-function variants. The American Journal of Human Genetics. 2021 Dec 2;108(12):2301–18. doi:10.1016/j.ajhg.2021.10.007 PubMed PMID: 34762822.

15. Landrum MJ, Lee JM, Riley GR, Jang W, Rubinstein WS, Church DM, et al. ClinVar: public archive of relationships among sequence variation and human phenotype. Nucleic Acids Res. 2014 Jan 1;42(D1):D980–5. doi:10.1093/nar/gkt1113

16. Mudge JM, Carbonell-Sala S, Diekhans M, Martinez JG, Hunt T, Jungreis I, et al. GENCODE 2025: reference gene annotation for human and mouse. Nucleic Acids Res. 2025 Jan 6;53(D1):D966–75. doi:10.1093/nar/gkae1078

17. Karolchik D, Hinrichs AS, Furey TS, Roskin KM, Sugnet CW, Haussler D, et al. The UCSC Table Browser data retrieval tool. Nucleic Acids Res. 2004 Jan 1;32(suppl_1):D493–6. doi:10.1093/nar/gkh103

18. Chang CC, Chow CC, Tellier LC, Vattikuti S, Purcell SM, Lee JJ. Second-generation PLINK: rising to the challenge of larger and richer datasets. Gigascience. 2015 Dec 1;4(1):s13742-015-0047–8. doi:10.1186/s13742-015-0047-8

19. Purcell S, Chang C. PLINK 2.0 [Internet]. Available from: www.cog-genomics.org/plink/2.0/

20. Koenig Z, Yohannes MT, Nkambule LL, Zhao X, Goodrich JK, Kim HA, et al. A harmonized public resource of deeply sequenced diverse human genomes. Genome Research. 2024 Jun 25;34(5):796–809. doi:10.1101/gr.278378.123 PubMed PMID: 38749656.

21. Bergström A, McCarthy SA, Hui R, Almarri MA, Ayub Q, Danecek P, et al. Insights into human genetic variation and population history from 929 diverse genomes. Science. 2020 Mar 20;367(6484):eaay5012. doi:10.1126/science.aay5012

22. The All of Us Research Program Investigators. The “All of Us” Research Program. The New England journal of medicine. 2019 Aug 15;381(7). doi:10.1056/NEJMsr1809937 PubMed PMID: 31412182.

23. Kars ME, Başak AN, Onat OE, Bilguvar K, Choi J, Itan Y, et al. The genetic structure of the Turkish population reveals high levels of variation and admixture. Proceedings of the National Academy of Sciences. 2021 Sep 7;118(36):e2026076118. doi:10.1073/pnas.2026076118

24. Naslavsky MS, Scliar MO, Yamamoto GL, Wang JYT, Zverinova S, Karp T, et al. Whole-genome sequencing of 1,171 elderly admixed individuals from Brazil. Nat Commun. 2022 Mar 4;13(1):1004. doi:10.1038/s41467-022-28648-3

25. Martin AR, Williams E, Foulger RE, Leigh S, Daugherty LC, Niblock O, et al. PanelApp crowdsources expert knowledge to establish consensus diagnostic gene panels. Nat Genet. 2019 Nov;51(11):1560–5. doi:10.1038/s41588-019-0528-2

26. Aartsma-Rus A. Overview on AON Design. In: Exon Skipping [Internet]. Humana Press; 2012 [cited 2026 Jul 10]. p. 117–29. Available from: https://link.springer.com/protocol/10.1007/978-1-61779-767-5_8 doi:10.1007/978-1-61779-767-5_8

27. Newton L, Haque B, Cheerie D, Tsoi CT, Klamann C, Sakaki R, et al. A scalable platform for exon-skipping antisense oligonucleotide therapy development for inborn genetic diseases [Internet]. medRxiv; 2026 [cited 2026 Aug 4]. p. 2026.07.29.26359175. Available from: https://www.medrxiv.org/content/10.64898/2026.07.29.26359175v1 doi:10.64898/2026.07.29.26359175

28. Guez J, Goodrich JK, Moldovan MA, Chao KR, Kar P, Panchal R, et al. Integrating 730,947 exome sequences with clinical literature improves gene discovery [Internet]. medRxiv; 2026 [cited 2026 Sep 3]. p. 2026.03.23.26349081. Available from: https://www.medrxiv.org/content/10.64898/2026.03.23.26349081v1 doi:10.64898/2026.03.23.26349081

29. D’Gama AM, Mulhern S, Sheidley BR, Boodhoo F, Buts S, Chandler NJ, et al. Evaluation of the feasibility, diagnostic yield, and clinical utility of rapid genome sequencing in infantile epilepsy (Gene-STEPS): an international, multicentre, pilot cohort study. The Lancet Neurology. 2023 Sep 1;22(9):812–25. doi:10.1016/S1474-4422(23)00246-6 PubMed PMID: 37596007.

30. Nguyen JNH, Lachgar-Ruiz M, Higginbotham EJ, Coleman M, Coleman J, Shao W, et al. Diagnostic Yield of Comprehensive Reanalysis After Nondiagnostic Short-Read Genome Sequencing in Infants With Unexplained Epilepsy. Neurology. 2026 Mar 24;106(6):e214645. doi:10.1212/WNL.0000000000214645

31. Helm J, Schöls L, Hauser S. Towards Personalized Allele-Specific Antisense Oligonucleotide Therapies for Toxic Gain-of-Function Neurodegenerative Diseases. Pharmaceutics. 2022 Aug;14(8):1708. doi:10.3390/pharmaceutics14081708

32. Clatot J, Ginn N, Costain G, Goldberg EM. A KCNC1 related neurological disorder due to gain of Kv3.1 function. Ann Clin Transl Neurol. 2022 Nov 23;10(1):111–7. doi:10.1002/acn3.51707 PubMed PMID: 36419348; PubMed Central PMCID: PMC9852383.

33. Jha V, Tsetsos C, Bedford M, Costain G, Vohra S, Diaz Martinez JP, et al. An n of 1 gene directed drug repurposing trial for an ultrarare genetic condition. Epilepsia. 2026 Jul;67(7):3644–56. doi:10.1002/epi.70253 PubMed PMID: 41999155; PubMed Central PMCID: PMC13360919.

34. Guillen Sacoto MJ, Tchasovnikarova IA, Torti E, Forster C, Andrew EH, Anselm I, et al. De Novo Variants in the ATPase Module of MORC2 Cause a Neurodevelopmental Disorder with Growth Retardation and Variable Craniofacial Dysmorphism. Am J Hum Genet. 2020 Aug 6;107(2):352–63. doi:10.1016/j.ajhg.2020.06.013 PubMed PMID: 32693025; PubMed Central PMCID: PMC7413887.

35. Gaviglio AM, Skinner MW, Lou LJ, Finkel RS, Augustine EF, Goldenberg AJ. Gene targeted therapies: Towards equitable development, diagnosis, and access. American J of Med Genetics Pt C. 2023 Mar;193(1):56–63. doi:10.1002/ajmg.c.32032

36. Giannikopoulos P, Lauffer MC, Marshall CR, Costain G, Niu Z, Bick D, et al. Interventional genomics: Bridging germline diagnosis and therapeutic action. Genetics in Medicine. 2026 Jun 1;28(6):102532. doi:10.1016/j.gim.2026.102532

37. Lee TL, Sawai T. Navigating equity in global access to genome therapy expanding access to potentially transformative therapies and benefiting those in need requires global policy changes. Front Genet. 2024 Apr 4;15:1381172. doi:10.3389/fgene.2024.1381172

38. Shickh S, Fooks K, Venkataramanan V, Acker M, MacDonald KV, Seeger TA, et al. A comprehensive approach to evaluating the clinical utility of genome sequencing in rare disease: A large prospective Canadian cohort. Genetics in Medicine. 2026 Apr 1;28(4):101684. doi:10.1016/j.gim.2026.101684

39. Greenfield A. Preclinical research (on rare diseases): we need to talk about health equity. Mamm Genome. 2025 Jun;36(2):347–53. doi:10.1007/s00335-024-10080-1

40. Smith JL, Adebamowo CA, Adebamowo SN, Darst BF, Fullerton SM, Gogarten SM, et al. Recommendations for responsible use of population descriptors in polygenic risk score development. Nat Genet. 2025 Dec;57(12):2962–71. doi:10.1038/s41588-025-02395-9

41. Appelbaum PS, Burke W, Parens E, Zeevi DA, Arbour L, Garrison NA, et al. Is there a way to reduce the inequity in variant interpretation on the basis of ancestry? Am J Hum Genet. 2022 Jun 2;109(6):981–8. doi:10.1016/j.ajhg.2022.04.012 PubMed PMID: 35659933; PubMed Central PMCID: PMC9247826.

42. Ying D, Cheung CL, O CK, Lam WKJ, Au Yeung SL, Lau CS, et al. Population-scale genomic medicine with the Hong Kong Genome Project. Nat Med. 2026 Jun;32(6):2277–87. doi:10.1038/s41591-026-04410-w

43. Finkel RS, Mercuri E, Darras BT, Connolly AM, Kuntz NL, Kirschner J, et al. Nusinersen versus Sham Control in Infantile-Onset Spinal Muscular Atrophy. New England Journal of Medicine. 2017 Nov 2;377(18):1723–32. doi:10.1056/NEJMoa1702752

44. gnomAD HGDP + 1KG callset [Internet]. [cited 2026 Jul 5]. Available from: https://gnomad.broadinstitute.org/data#v3-hgdp-1kg

45. Costain Lab GitHub [Internet]. Available from: github.com/CostainLab/knockdown_aso

