## Supplemental Material 2 for "Population-scale genome-wide prioritization of targets for allele-specific antisense oligonucleotide therapy design"

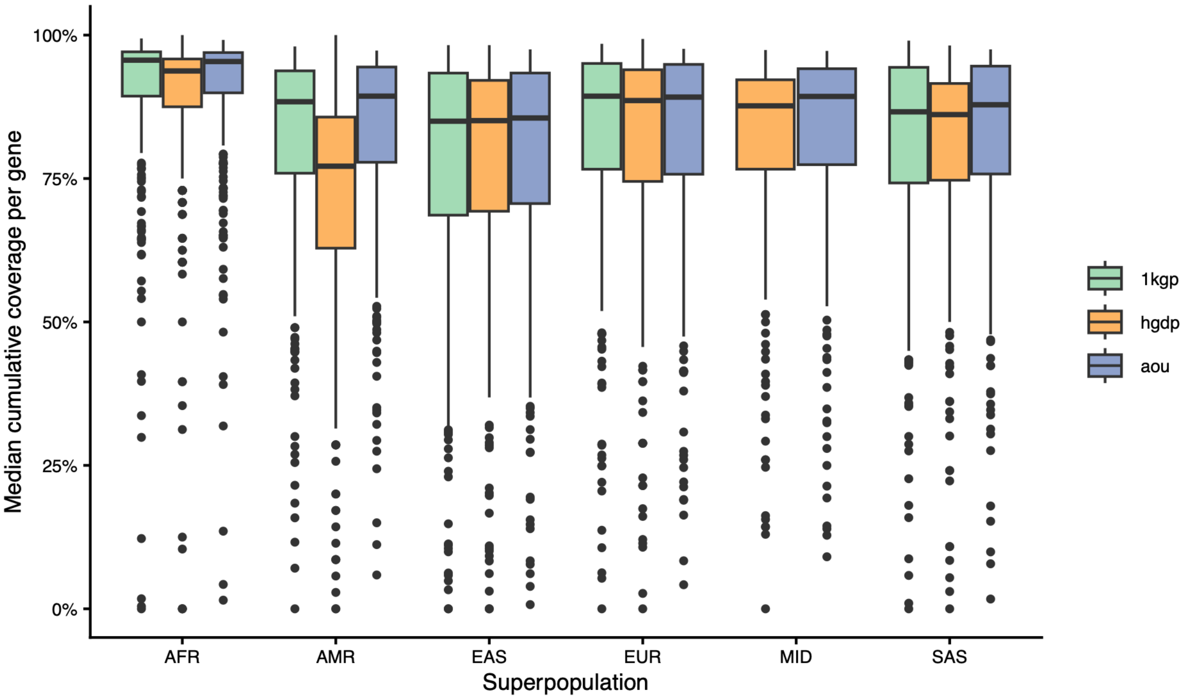


Supplemental Fig. 1: Boxplot showing the distribution of cumulative heterozygous population coverage across the 1000 Genomes Project, the Human Genome Diversity Project, and the *All of Us* Research Program variant call datasets. Boxes represent the interquartile range (IQR), the central line indicates the median, whiskers extend to observations within 1.5 × the IQR and points represent outliers. AFR, African; AMR, Admixed American; EAS, East Asian; EUR, European; MID, Middle Eastern; SAS, South Asian.
